# Optimizing Genetic Testing Strategy for Suspected Attenuated Adenomatous Polyposis: Effective Solutions in Public Health Systems

**DOI:** 10.1101/2024.06.28.24308416

**Authors:** Natalia García-Simón, Fátima Valentín, Ana Royuela, Beatriz Hidalgo-Calero, Ricardo Blázquez-Martín, Montserrat de-Miguel-Reyes, José M Sánchez-Zapardiel, Luisa Adán-Merino, Alejandro Rodríguez-Festa, Patricia Gallego-Gil, Pilar Mediavilla-Medel, Laura Quiñonero-Moreno, Lourdes Gutiérrez, Alberto Herreros-de-Tejada, Antonio Sánchez, Mariano Provencio, Atocha Romero

## Abstract

**BACKGROUND:** Attenuated familial adenomatous polyposis (AFAP) and MUTYH-associated polyposis (MAP) represent key hereditary attenuated adenomatous polyposis syndromes, with *APC* and *MUTYH* being the associated genes. While guidelines recommend genetic testing based on polyp count for diagnosing these syndromes, age is often overlooked despite its influence on polyp prevalence.

**AIM:** To enhance genetic testing strategies for suspected attenuated adenomatous polyposis by combining polyp count and age in a probability calculator.

**METHODS:** Retrospective study of adult patients referred to NGS genetic testing for suspected attenuated adenomatous polyposis (accumulated history of <100 adenomas) (discovery cohort, N=138). Data collected included age, adenoma count at the time of genetic testing, and test results. Multivariable logistic regression model was developed to link a positive genetic test result with age and adenoma count. The model was externally validated with populations from two tertiary hospitals in our region (validation cohort, N=259).

**RESULTS:** In the discovery cohort, 13 (9.4%) patients tested positive for pathogenic mutations. Positive cases were younger (odds ratio (OR): 0.91, 95% confidence interval (95%IC): 0.86-0.96) and developed more adenomas (OR: 1.08, 95%IC: 1.04-1.13) compared to negative cases. The logistic regression model combining age and polyp count demonstrated an AUC of 0.92. Using a cutoff probability of 3.5%, the model achieved 100% sensitivity and 58% specificity in identifying positive cases. In the external validation, the model accurately predicted 14 out of 16 positive cases (88%). The remaining two positive cases were a patient with an *AXIN2* mutation in heterozygosis, and a patient with a *NTHL1* mutation in homozygosis. Performance evaluation of both hospitals yielded AUC values of 0.77 and 0.90.

**CONCLUSIONS:** Older individuals, particularly those with few polyps, may have diminished chances of detecting hereditary syndromes. Incorporating age as a criterion for genetic testing referral has the potential to refine patient selection and improve cost-effectiveness.

**Core tip:** This retrospective muli-center observational study aims to investigate the weight of age and polyp count on patients diagnosed with hereditary attenuated adenomatous polyposis syndrome compared to those with sporadic polyposis. Base on this, a probability calculator has been developed to enhance the efficiency and cost-effectiveness of genetic testing protocols for attenuated adenomatous polyposis syndromes within public health systems.

## INTRODUCTION

Hereditary polyposis syndromes are known to be accountable for about 2-3% of all cases of colorectal cancer (CRC)^[1–3]^. The most common polyposis syndromes are familial adenomatous polyposis (FAP) (OMIM #175100), attenuated FAP (AFAP) (OMIM #175100), and *MUTYH*-associated polyposis (MAP) (OMIM #608456), while other syndromes such as hamartomatous polyposis are less frequent^4,5^. The main genes associated with hereditary adenomatous polyposis syndromes are *APC* (OMIM #611731) gene, for FAP and AFAP, and *MUTYH* (OMIM #604933) gene, for MAP.

For suspected patients, guidelines recommend offering genetic testing based on the number of polyps, with a threshold of more than 100 adenomatous polyps for FAP, and more than 10 or 20 adenomatous polyps (depending on the guideline) for AFAP and MAP^[6–10]^. However, since polyps are not only caused by mutations in polyposis genes but are also intrinsic to age, the older the patient is, the more likely it is to detect polyps, lowering the probability of being a case of hereditary syndrome, especially when the polyp burden is low. Therefore, despite the selection of patients, germline multigene testing continues to have a high demand in laboratories, which decreases the rate of mutation detection, making these studies not cost-effective. Stanich *et al.*^[11]^ demonstrated that, on the one hand, the prevalence of mutations in adenomatous polyposis syndromes genes (*APC* and *MUTYH*) increases with the number of polyps developed, and on the other hand, older populations have a lower prevalence of finding significant mutations in these genes. More specifically, in those patients with a low number of polyps (<20), this prevalence is reduced below 2% from 50 years and onwards.

Consequently, age should also be included as a criterion for referring to genetic testing, helping the selection of patients, although very few guidelines include it. In this paper, we aim to improve genetic testing performance in suspected attenuated adenomatous polyposis by establishing a probability calculator based on the number of polyps and age upon which recommend referring to genetic testing.

## METHODS

### Subjects

We conducted a retrospective analysis of all patients aged 18 years and older who were referred for genetic testing at Puerta de Hierro Hospital due to suspicion of attenuated adenomatous polyposis (AFAP or MAP) between 2015 and 2023 (N=138). The suspicion of attenuated polyposis was based on an accumulated history of 10 to 100 adenomatous polyps, according to the Community of Madrid (CAM) guidelines followed by our center^12^. Patients with two or more hamartomatous polyps were excluded from the study, regardless of the number of adenomas. The presence of hamartomatous polyps suggests hamartomatous polyposis and excludes adenomatous polyposis^6^. The study received approval from the ethics committee of Puerta de Hierro Hospital (internal code: PI_48/24). Pre-test genetic counseling was conducted, and clinical consent for genetic testing was obtained.

Only pathogenic (P) (class 5) and likely pathogenic (LP) (class 4) variants in genes related to hereditary adenomatous polyposis (*APC* and *MUTYH*) were considered positive cases. Being a recessive gene, *MUTYH* variants were classified as positive cases only if found in homozygosity or compound heterozygosity. No variants detected, benign (class 1) and probably benign (class 2) variants, or monoallelic variants in *MUTYH* gene, were all considered negative cases.

### Genetic testing

At Puerta de Hierro Hospital, germline DNA was extracted from peripheral blood using the Maxwell RSC whole blood DNA kit (Promega). Genetic testing was performed by massive sequencing (NGS) on a (MiSeq sequencer (Illumina) using the Hereditary Cancer Solution (HCS) kit (Sophia Genetics) and following the manufacturer’s instructions. The panel included *APC* and *MUTYH* as relevant genes associated with adenomatous polyposis. Bioinformatic analysis was performed using the Sophia DDM-V4 (Sophia Genetics) data analysis platform. Relevant SNPs and indels were confirmed by Sanger sequencing. The reference sequences used to name variants were NM_001128425.2 for *MUTYH* and NM_000038.6 for *APC*.

### Age and number of polyps

Age data is referred to as years of age at the moment of the genetic testing.

Number of polyps refers to the total accumulative polyps until genetic testing. Information related to colonoscopy examinations was collected with ENDOBSE® (Olympus Corporation, Tokyo, Japan). The program reflected all the data from the colonic examination: quality of the preparation, type of sedation, number of polyps, and location, along with other patient data relevant to the examination such as reason for endoscopic study and medication.

Histologically, polyps were classified into the following groups: adenomatous polyps, sub-classified in tubular, tubulovillous and villous, and non-adenomatous polyps, sub-classified in hyperplastic and serrated polyps. There were some reports that describe resected polyps just as “adenomatous” with no sub-classification, so they are here reported as “not classified” adenomatous polyps and were only considered in the adenomatous vs non-adenomatous polyps comparison and not in the subtype comparison.

### External validation

In order to validate the findings, we utilized two independent cohort datasets (N=259). The datasets were obtained from 12 de Octubre University Hospital (n=162) and Infanta Leonor University Hospital (n=97).

As for methodology, the 12 de Octubre University Hospital, extracted DNA from whole blood using the Maxwell RSC Whole Blood kit (Promega). The Custom Hereditary Cancer Solution (CHCS) kit (Sophia Genetics) was employed for genetic testing, and software analysis was conducted using Sophia DDM-V4 (Sophia Genetics). Genes included in the sequencing kit were *APC, MUTYH, POLE, POLD1, AXN2* and *NTHL1*. Any pathogenic or likely pathogenic variants identified through massive sequencing were subsequently validated via Sanger sequencing.

Infanta Leonor Hospital utilized the QIAamp Blood DNA kit (QIAcube) for the extraction and purification of DNA from peripheral blood. Genetic testing was conducted by NGS on a MiSeq (Illumina) using the SureSelect QXT Target Enrichment (Agilent) kit for the coding region and flanking zones of the analyzed genes (*APC, MUTYH, POLE, POLD1, NTHL1, MSH3*). The bioinformatic analysis was carried out using custom-designed analysis pipelines, assisted by the SureCall and Alissa Interpreter software (Agilent). Sanger sequencing was employed to confirm relevant SNPs.

### Statistics

The Shapiro-Wilk test was employed to assess normality. Non-normally distributed quantitative variables were presented as median along with the 25th (P25) and 75th (P75) percentiles. For nonparametric comparisons, the Chi square test and Mann-Whitney test were used for categorical and quantitative variables respectively. Multivariable logistic regression (logit) was used to establish the association between having a positive result in the genetic test (dependent variable) and the age and number of adenomatous polyps. The model was internally validated through the bsvalidation command in Stata^[13]^. This command performs an internal validation through calibration and discrimination. Resampling techniques were performed by bootstrapping, with 500 replications. To evaluate calibration, a calibration plot was generated, in which the quintiles of the observed and expected probabilities of having the event were graphically confronted. If calibration is perfect, the line between the two risks will lie on the main diagonal of the plot. The expected/observed (E/O) ratio will equal 1, the calibration in the large (CITL) will be 0 and the slope equal to 1. Discrimination is measured by the C-statistic, which is an analog of the AUC, with values ranging from 0.5 for no discrimination to 1.0 for perfect discrimination. The Brier scale (range 0-100) was also calculated as an overall performance measure, with high values indicating predictions closer to the actual outcome. It was obtained from the Brier score: Brier scaled = 1 – Brier score / Brier max.

For the external validation, the calibration plot assessed the calibration and the C-statistic, the discrimination. The Brier score is also shown, with a range between 0 and 1, being the lower value, the more accurate prediction.

From the model predicted probability, we pursued an optimal cutoff point with the maximal sensitivity and developed an online calculator available at https://investigacionpuertadehierro.com/laboratorio-biopsia-liquida/.

P value <0.05 was considered statistically significant.

Statistical analysis was carried out using MedCalc Statistical Software version 11.4.2.0 program (MedCalc Software bvba, Ostend, Belgium; http://www.medcalc.org; 2018), Stata v18 (StataCorp. 2023. *Stata Statistical Software: Release 18*. College Station, TX: StataCorp LLC.).

## RESULTS

Of the total of patients included in the Puerta de Hierro cohort (N=138), 13 patients (9.4%) had a positive result in the genetic testing. From these, 11 patients had a P/LP variant in *MUTYH* gene, of which three were in homozygosis and eight in compound heterozygosis; two patients had a P variant in *APC* gene in heterozygosis (Supplementary figure 1). The most prevalent *MUTYH* mutation was c.1187G>A p.(Gly396Asp) (commonly known as G396D), present in the three homozygous cases and half of compound heterozygous cases, followed by c.536A>G p.(Tyr179Cys) (commonly known as Y179C) mutation found in the other half of compound heterozygous cases. Other mutations identified in this biallelic group were c.1012C>T p.(Gln338Ter) in 3/8 cases, c.1227_1228dup p.(Glu410fs) in 2/8 cases, and c.933+3A>C, c.736G>T p.(Val246Phe) and c.1101dup in 1/8 each one. Table 1 shows detailed information on the positive cases.

**Table 1.** Characteristics of positive group patients. GT: genetic testing; CRC: colorectal cancer; FH: family history. * The exact age data is not shown due to editorial request; it will be available upon journal publication.

| Patient | Range age at GT (years)* | Gene | Variant | Heterozygosity | CRC (Range age diagnosis)* | FH CRC |
| --- | --- | --- | --- | --- | --- | --- |
| **Case 1** | 70-75 | *MUTYH* | c.1187G>A | Homozygous | YES (55-60) | NO |
| **Case 2** | 25-30 | *MUTYH* | c.536A>G + c.933+3A>C | Compound heterozygous | NO | YES |
| **Case 3** | 45-50 | *MUTYH* | c.1012C>T + c.536A>G | Compound heterozygous | NO | YES |
| **Case 4** | 40-45 | *MUTYH* | c.1012C>T + c.536A>G | Compound heterozygous | NO | YES |
| **Case 5** | 45-50 | *MUTYH* | c.1012C>T + c.536A>G | Compound heterozygous | NO | YES |
| **Case 6** | 60-65 | *MUTYH* | c.1187G>A + c.736G>T | Compound heterozygous | YES (55-60) | YES |
| **Case 7** | 65-70 | *MUTYH* | c.1187G>A | Homozygous | YES (55-60) | YES |
| **Case 8** | 75-80 | *MUTYH* | c.1187G>A | Homozygous | NO | YES |
| **Case 9** | 50-55 | *MUTYH* | c.1187G >A + c.1227_1228dup | Compound heterozygous | NO | NO |
| **Case 10** | 50-55 | *MUTYH* | c.1187G >A + c.1101dup | Compound heterozygous | NO | NO |
| **Case 11** | 40-45 | *MUTYH* | c.1187G >A +<br>c.1227_1228dup | Compound heterozygous | YES (40-45) | NO |
| **Case 12** | 35-40 | *APC* | c.697C>T | Heterozygous | NO | YES |
| **Case 13** | 75-80 | *APC* | c.423G >C | Heterozygous | NO | NO |

Patient characteristics of positive and negative groups are shown in Table 2. According to sex, there were no statistical differences (odds ratio (OR): 0.91, 95% confidence interval (95%IC): 0.86 to 0.96, *P =* 0.012). Proportionally, development of CRC was similar between the two groups, with 4 (24.8%) CRC patients in the positive group and 31 (30.8%) in the negative group (OR: 1.35, 95%CI: 0.39 to 4.69, *P =* 0.64). Regarding family history (FH) of CRC, 43.2% of patients in the negative group had at least one family member with CRC while for the positive group, the percentage rose to 61.5%, although the difference did not reach statistical significance (OR: 2.04, 95%IC: 0.63 to 6.57, *P =* 0.23).

**Table 2.** Patients’ characteristic for positive and negative group. CRC: colorectal cancer; FH CRC: family history of colorectal cancer; ND: no data.

| Patients' characteristics | Positive group<br>n=13 | Negative group<br>n=125 | P value |
| --- | --- | --- | --- |
| Age (years)<br>median (P25-P75) | 51 (44 - 66) | 67 (61 - 72) | 0.012 |
| Sex |  |  | 0.51 |
| Women, n (%) | 6 (46.2) | 46 (36.8) |  |
| Men, n (%) | 7 (53.8) | 79 (63.2) |  |
| Polyp type | 701 (100) | 3685 (100) | <0.001 |
| Adenomatous, n (%) | 677 (96.6) | 2916 (79.1) |  |
| Non-adenomatous, n (%) | 24 (3.4) | 769 (20.9) |  |
| CRC, n (%) | 4 (30.8) | 31 (24.8) | 0.64 |
| FH CRC, n (%) | 8 (61.5) | 55 (44) | 0.23 |
| Smoking, n (%) |  |  | 0.004 |
| Yes | 2 (16.6) | 42 (33.3) |  |
| Former | 1 (5.6) | 49 (39.7) |  |
| No | 7 (55.6) | 26 (20.6) |  |
| ND | 3 (22.2) | 8 (6.4) |  |

Parameters that did show significant differences between the negative and positive groups were age (OR: 0.91, 95%CI: 0.86 to 0.96, *P =* 0.012), adenoma number (OR: 1.08, 95%IC: 1.04 to 1.13, *P <* 0.001) and smoking (OR: 8.17, 95%IC: 1.97 to 33.8, *P =* 0.004).

### Age comparison study

The youngest positive case was 28 years old, and the oldest one was 79 years old. For the negative group, ages ranged from 39 to 83 years old. The median age was 51 years in the positive group (percentile25-P75: 44 to 66), whereas the median age was 67 years (P25-P75: 61 to 72) in negative cases (OR: 0.91, 95%CI: 0.86 to 0.96; Figures 1 and 2). Among all negative cases, 74% were aged over 60, whereas the positive group had only five cases above that age.

**Figure 1.**
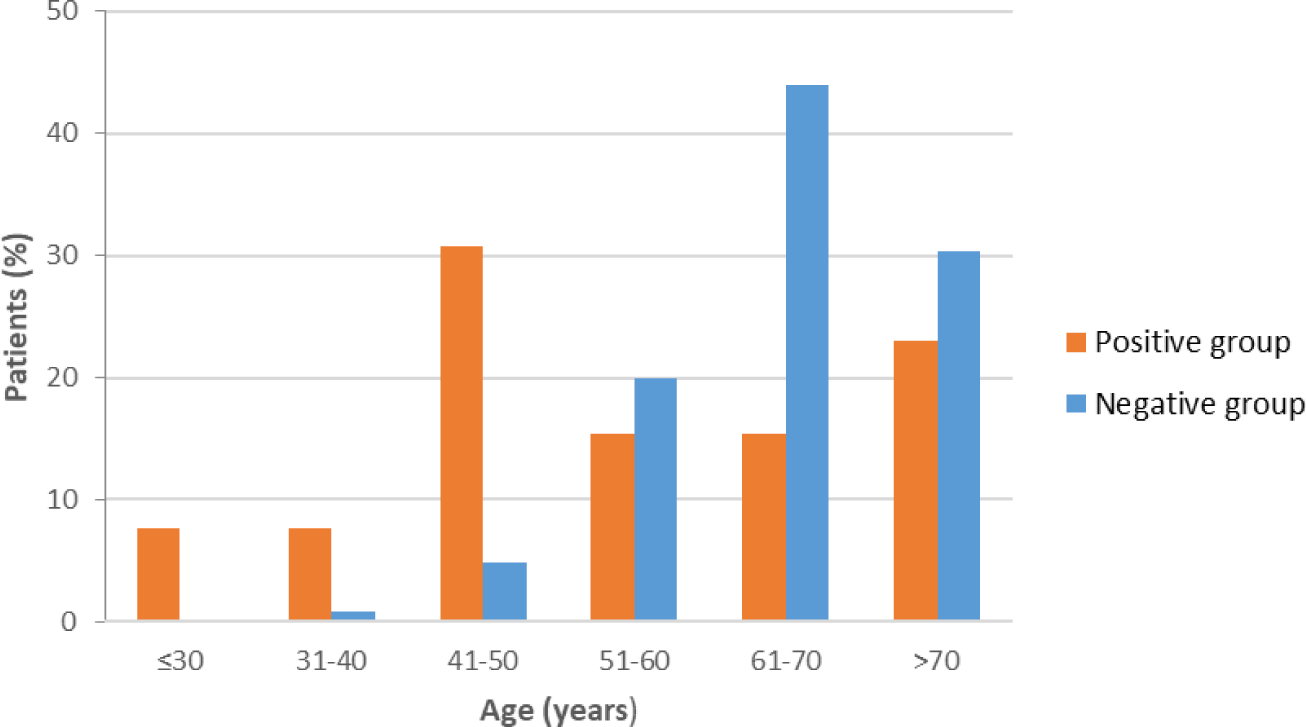
Distribution of ages (by decades of years) at which genetic testing was performed.

**Figure 2.**
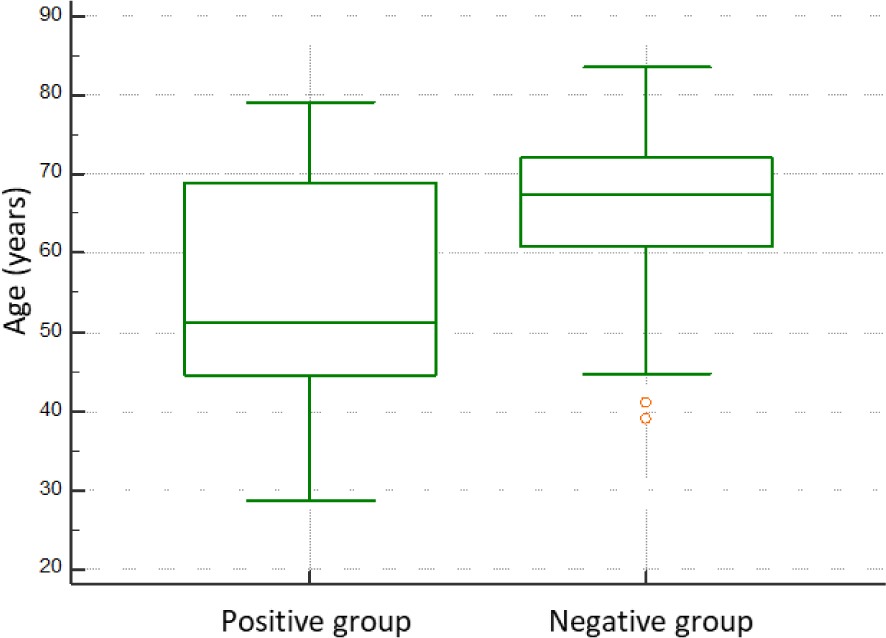
Box and whiskers plot of age at genetic testing for negative and positive.

### Polyp comparison study

As previously mentioned, comparison data reviled a relation between the genetic test result and the number of adenomas. Both groups developed more adenomatous polyps than non-adenomatous polyps, but overall, the positive group developed significantly more adenomas (median: 35, P25-P75: 32 to 74) than the negative group (median: 22, P25-P75: 16 to 28) (OR: 1.08, 95%IC; 1.04 to 1.13) (Table 3). At the time of genetic testing, the majority of positive cases (85%) accumulated more than 30 adenomas, while only 24% of negative cases reached that threshold (Figure 3).

**Figure 3.**
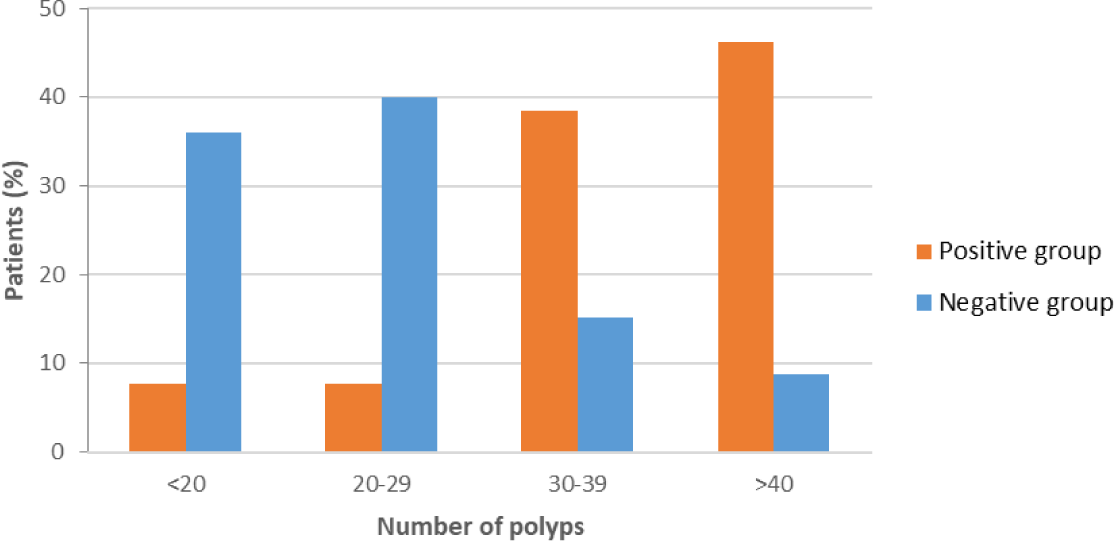
Distribution of number of adenomatous polyps by group.

**Table 3.** Distribution of adenomatous and non-adenoamtous polyps by subtype. *P* value was calculated comparing only tubular, tubulovillous and villous polyps, leaving out the not sub-classified polyps.

| Polyp subtype | Positive group<br>n=701 | Negative<br>group<br>n=3685 | P value |
| --- | --- | --- | --- |
| Adenomatous,<br>n (%) | 677 (96.6) | 2916 (79.1) | 0.42 |
| Tubular | 616 (91) | 2620 (89.9) |  |
| Tubulovillous | 35 (5.2) | 179 (6.1) |  |
| Villous | 3 (0.4) | 7 (0.2) |  |
| Not sub-classified | 23 (3.4) | 110 (3.8) |  |
| Non-adenomatous,<br>n (%) | 24 (3.4) | 769 (20.9) | 0.81 |
| Hyperplastic | 20 (83) | 626 (81) |  |
| Serrated | 4 (17) | 143 (19) |  |

There were no significant differences according to the subtypes of polyps. For adenomatous subtypes, the most common one was tubular in the positive group as well as in the negative group (91% vs 89.9% respectively), followed far behind by tubulovillous (5.2% vs 6.1%) and villous polyp subtypes (0.4% vs 0.2%) (*P =* 0.42). For non-adenomatous polyps, the hyperplasic subtype was the most prevalent (83% vs 81%) and serrated polyps were less frequent in both groups (17% vs 19%). The graphic distribution of polyps is represented in Supplementary figure 2.

### Calculator

#### Model development

Once all data was gathered, we aimed to establish a mathematical relation between the combination of age and number of adenomatous polyps, and the genetic test result. Using multivariable logistic regression, we estimated the probability of a patient having a positive genetic result based on their age and number of adenomas at genetic testing. The regression equation was: ***logit (genetic test (+)/1-genetic test (+)) = 0,3822 + (-0,0814*age in years) + (0,0731*number of adenomas)*** ^©^ IDIPHIM, (2024), All rights reserved. Overall model performance was ranked by a Brier score of 32.6%. Calibration scores were 1 for E/O ratio, 0 (95%CI: -0.73 to 0.73) for CITL, and 1 (95%CI: 0.56 to 1.44) for slope. Discrimination was assessed by an AUC of 0.924 (95%CI: 0.85 to 0.99; *P <* 0.01) (Figure 4).

**Figure 4.**
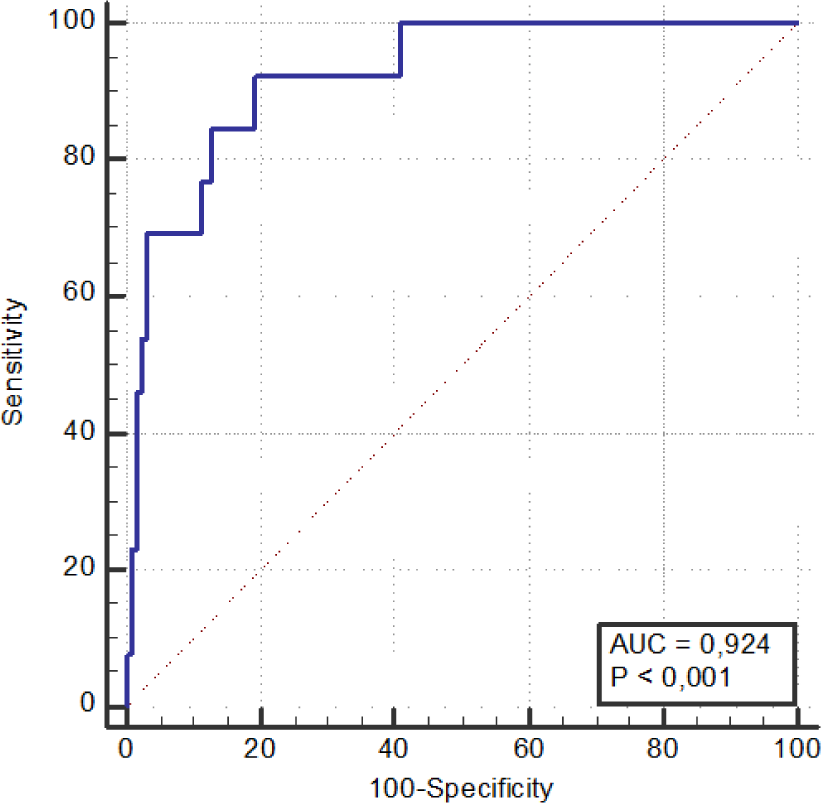
AUC for probability of having a positive genetic test.

The next step was to establish a cut-off point from the predicted probability model upon which to decide whether to refer patients to genetic testing or not. The requirement set to select this point was having a 100% sensibility with maximum specificity, so the false negative rate would be 0% but minimizing the number of false positives. These criteria were fulfilled at a probability of 3.5%, with a sensibility of 100% and a specificity of 58%. Applying the model retrospectively, it was found that 74 cases meeting the polyposis criteria of the CAM had a probability of a positive genetic test below 3.5%. This could have potentially led to savings amounting to 50,000€.

#### Internal validation

The internal validation was made by bootstrap resampling. The Brier score was 24.3% for overall model performance. In terms of calibration, the results obtained were an E/O ratio of 0.97 (95% CI: 0.57 to 1.38), CITL of 0.07 (95%CI: -0.8 to 1.01), and a calibration slope of 0.89 (95%CI: 0.39 to 1.51). C-statistic for discrimination was 0.9 (95%CI: 0.78 to 1). After the adjustment of the model by bootstrapping, the OR for age was 0.93 (95%CI: 0.88 to 0.98), and the OR for number of polyps was 1.07 (95%CI: 1.03 to 1.1).

#### External validation

The final validation was made by using data from other centers (N=259), located in the same geographical area. We gathered data on the number of polyps and age at the time of genetic testing, and the results of such test, classifying patients between “positive” (when genetic results revealed a mutation in genes related to polyposis) and “negative” (when no mutation related to polyposis was found). The general patient characteristics from external centers closely resembled those of our own, with a higher proportion of men than women and comparable median age and number of polyps (Table 4).

**Table 4.** Patients’ characteristics for study center and external validation centers.

| Patients<br>characteristics | Puerta de Hierro<br>(N=138) | 12 de Octubre<br>(n=162) | Infanta Leonor<br>(n=97) |
| --- | --- | --- | --- |
| Sex |  |  |  |
| Women, n (%) | 52 (37.7) | 43 (26.5) | 23 (23.7) |
| Men, n (%) | 86 (62.3) | 119 (73.5) | 74 (76.3) |
| Age,<br>median (P25-P75) | 67 (60-72) | 66 (59-71) | 67 (60-73) |
| Nº of polyps,<br>median (P25-P75) | 23 (16-31) | 21 (15-29) | 24 (19.8-32) |

At 12 de Octubre Hospital (n=162), 11 patients were reported as positive. Of these, 4 cases presented *MUTYH* mutations (3 homozygous and 1 compound heterozygous), 4 cases carried *APC* mutations in heterozygosis and 1 case presented a heterozygous *POLD1* mutation. The model correctly predicted the positive result in 9 out of these 11 cases. The remaining two positive cases were predicted as negative. One patient, in their late 60s, had 19 adenomatous polyps and a heterozygous *AXIN2* gene mutation c.1994dup p.(Asn666fs). The second patient, in their early 70s, had 20 adenomatous polyps a homozygous *NTHL1* gene mutation c.268C>T p.(Gln90Ter). Calibration performance yielded a Hosmer-Lemeshow p-value of 0.45 (Table 5), while the discrimination study resulted in an AUC of 0.77 (95%CI: 0.61 to 0.93) (Figure 5).

**Figure 5.**
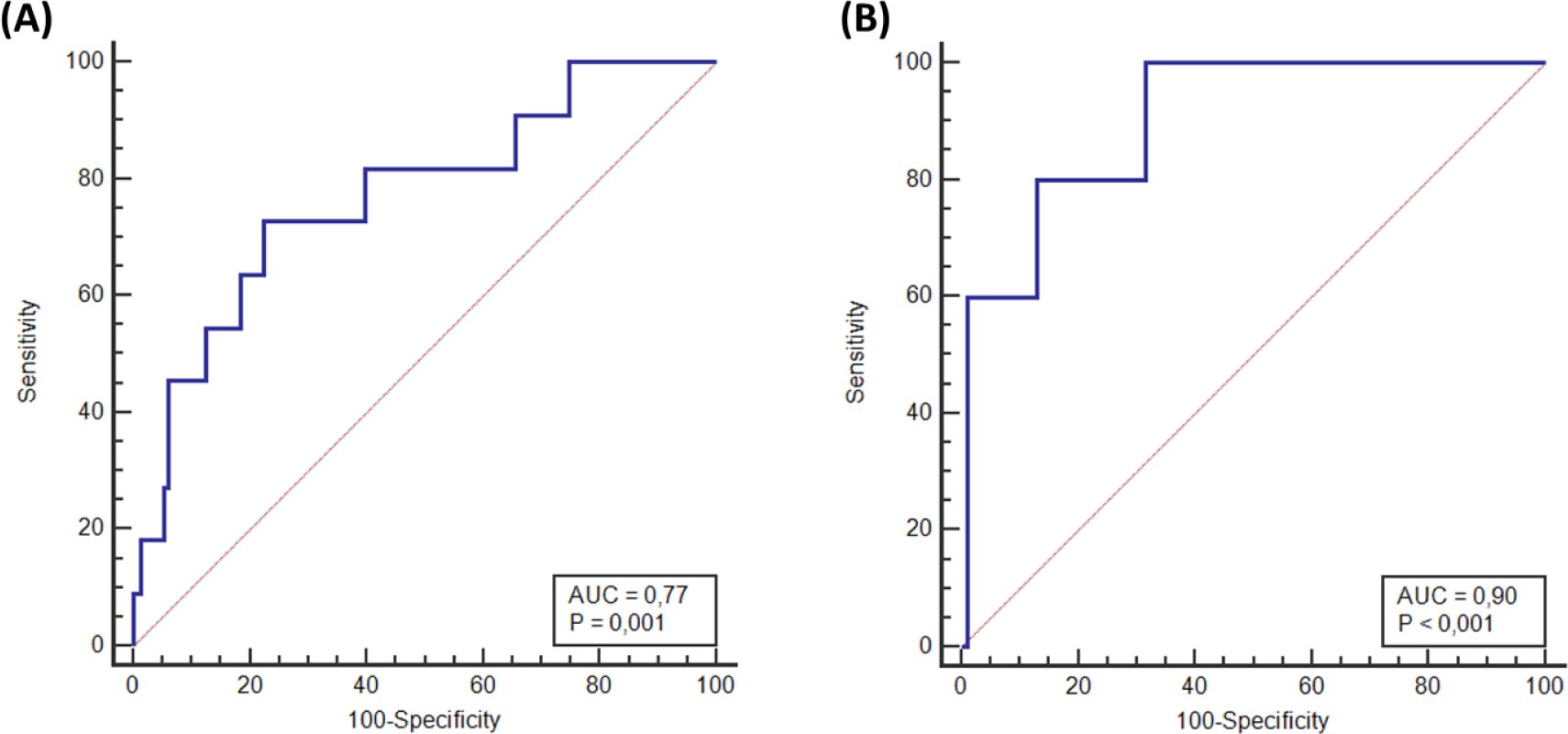
AUC resulting from applying the model to external validation databases. (A) AUC for 12 de Octubre Hospital, and (B) AUC for Infanta Leonor Hospital

**Table 5.** Perform results for external validation data.

| Hospital | Patients |  |  | Model prediction |  | Performance measures |  |
| --- | --- | --- | --- | --- | --- | --- | --- |
|  | All<br>n | Positive<br>n, (%) | Negative<br>n, (%) | TP<br>n, (%) | TN<br>n, (%) | Hosmer-Lemeshow p-value | AUC |
| 12 de Octubre | 162 | 11 (6.8) | 151 (93.2) | 9 (81.8) | 75 (49.7) | 0.45 | 0.77 |
| Infanta Leonor | 97 | 5 (5.2) | 92 (94.8) | 5 (100) | 49 (53.3) | 0.38 | 0.90 |

At Infanta Leonor Hospital (n=97), 92 cases were negative, and 5 cases were positive. Mutations identified in these patients included three in *MUTYH* (1 homozygous and 2 compound heterozygous), one in *APC,* and one in *PTEN*. All 5 mutated patients were accurately predicted as positive. Performance evaluation indicated a Hosmer-Lemeshow p-value of 0.38 (Table 5) and an AUC of 0.90 (95%CI: 0.78 to 1) (Figure 5).

## DISCUSSION

The diagnosis of Hereditary Polyposis Syndromes is of utmost importance due to the implications for not only patients but also their family relatives. This diagnosis begins with an oligopoliposis phenotype, and it is confirmed by genetic testing when a mutation in one of the implicated genes is found. Ensuring an accurate selection of patients for genetic testing is indispensable for the efficiency of public health systems. Since polyps are not only caused by genetic mutations but are also caused by aging, there is a phenotype overlap between hereditary polyposis and sporadic polyposis. The CAM implemented the *Prevecolon* program in 2017 for the early diagnosis of CRC through the fecal occult blood test (FOBT)^[14]^. The campaign involves mass screening of asymptomatic individuals aged 50 to 69, who undergo the FOBT, and if positive, are referred for colonoscopy. This campaign has resulted in an increase in the detection of asymptomatic polyposis, leading to a greater number of patients being referred for genetic consultation and consequently resulting in higher expenditure of both material and human resources^[15–18]^. Therefore, this translates into a high rate of negative results, and thus, a low diagnostic yield. All of this emphasizes the need to implement new tools for better patient selection. To improve it, guidelines include other features such as personal or family history of CRC to help a better distinction between genetic and sporadic polyposis^[19–21]^. Nowadays age is beginning to be included too, although very few guidelines do it and there is no consensus about the cut-off limit^[6,9,22]^. Consequently, we sought to identify differences between hereditary and sporadic polyposis in order to improve the selection process. Out of that, we created a calculator based on the number of adenomas and age to help the health professionals make a better selection of patients and optimize the diagnostic yield of genetic testing.

In the Puerta de Hierro cohort, the prevalence of biallelic *MUTYH* mutations was 8% (11/138), and 1.5% (2/138) for *APC* mutations, in line with previous studies which ranged prevalence of these mutations in patients with oligopolyposis from 3% to 15% for *MUTYH* and from 2% to 9% for *APC*^[23–27]^.

Among the *MUTYH* pathogenic variants found, the most represented ones were G396D and Y179C. This is consistent with what has been previously found since most patients belonged to European population, in which these two variants are considered founder mutations^[24,28–31]^. In the case of *APC* mutations, it has been reported that phenotype arising from alterations in this gene, varies depending on the location of the mutation. The ones found in this study fall into the regions of 5’ end (codons 1 to 233) and exon 9 (codons 311 to 412), which have been associated with AFAP^[7,32–35]^.

Confrontation of other features between positive and negative group, showed no differences in sex, as described in other studies^[36–38]^. Personal history of CRC and family history of CRC did not reach statistical signification between the two groups, demonstrating that the CRC risk for mutated patients in this study has been lowered due to the early diagnosis and prophylactic surgical strategies carried out (polypectomies and colectomies) that prevented developing CRC^[39–43]^.

In terms of tobacco consumption, the negative group exhibited higher rates of smoking and former smoking compared to the positive group (OR: 8.17, 95%IC: 1.97 to 33.8). Tobacco is a known carcinogen and has been linked to the development of polyposis^[44–46]^. Our data imply that smoking was a significant contributing factor in sporadic polyposis cases.

Among the different polyps that can be present in general in polyposis, adenomatous polyps represent about two-thirds of all colonic polyps (being tubular adenomas the major representative of this group) followed by hyperplasic polyps as the second most common polyps. Tubulovillous and villous adenomas, as well as serrated polyps, can also be normally found but are less frequent^[38,47]^. In this study, mutated patients developed in proportion more adenomas and much less hyperplasic polyps than not mutated cases. Interestingly, if the comparison was made separately among adenomatous subtypes on one side, and non-adenomatous subtypes on the other, there were no differences in the distribution. For both positive and negative groups, tubular polyp histology was the most common one among the adenomatous polyps, followed by tubulovillous polyps and villous polyps, while 80% of non-adenomatous polyps had hyperplasic histology. This demonstrates that altered genomics has more penetrance than sporadic predisposing factors (such as age) for the development of adenomatous polyps. However, mutations in polyposis-related genes do not influence the distribution of subtypes.

For age comparison, the median age for patients with an *APC* or biallelic *MUTYH* mutation was 51 years, significantly younger than those patients in the negative group (OR: 0.91, 95%CI: 0.86 to 0.96). Our results complement other studies that reached the same conclusion^[11,32,48–50]^. This consolidates the use of age as a complementary criterion for referring to genetic test. In sum, our data stated that hereditary polyposis patients develop polyps at an earlier age and with a higher proportion of adenomas.

Using data on the number of adenomas, age, and genetic test results, we constructed a model to estimate the likelihood of detecting a polyposis mutation based on adenoma count and age. After evaluating various thresholds, we established the decision point at a 3.5% probability, ensuring 100% sensitivity and nearly 60% specificity. Had this model been applied to the patients of the Puerta de Hierro Hospital cohort, more than half of the genetic tests (53.6%) could have been saved, avoiding any missed positive cases and resulting in savings of 50,000€. External validation was conducted using data from two different hospitals within the same geographical area. This approach aimed to minimize potential confounding factors associated with variations in patient characteristics. Performance evaluation at both centers reported a Hosmer-Lemeshow p-value above 0.05, indicating no significant differences between observed and model-predicted values. In terms of discrimination, the area under the curve (AUC) was satisfactory for both centers. However, 12 de Octubre Hospital exhibited slightly poorer performance due to two positive cases being incorrectly predicted as negative by the model. Both cases involved elderly patients (over 65 years) with a low number of polyps (19 and 20 adenomas). Guidelines^6,8,9,10^ are gradually shifting the adenoma count threshold for recommending genetic testing from 10 to 20, and those that include age, criteria are more restricted for patients over 60 years old. These two cases fell into a grey area, as one was below the 20-adenoma threshold and the other one was just in the limit with an advanced age. Consequently, depending on the guidelines applied, these two patients might not have met the requested criteria for genetic testing referral.

Our study has some limitations. The model is constructed on the basis of solely common adenomatous polyposis genes, *APC* and *MUTYH*, as positive cases. However, in recent years, new genes such as *POLE, POLD1, AXIN2,* and *NTHL1* have been associated with adenomatous polyposis. These genes are being incorporated into genetic panels but were not available at the time of the testing in our center, and therefore, were not included in the model. However, given that the phenotype is very common among the different genes, this limitation may not be significant. The model correctly predicted a *POLD1* carrier, while the *AXIN2* and *NTHL1* cases that were wrongly predicted were on the limit of the cutoff point and could potentially have been correctly predicted with data from the next colonoscopies.

## CONCLUSION

In conclusion, hereditary polyposis syndromes present themselves at an early age and with a higher burden of adenomas than sporadic polyposis. Both features should be taken into consideration for selecting patients to refer to genetic testing. To ease the process, we developed a calculator that provides the probability of obtaining an informative genetic result based on these two characteristics. This will aid in deciding whether to proceed with genetic testing.

## Data Availability

All data produced in the present study are available upon reasonable request to the authors

https://investigacionpuertadehierro.com/calculadora-poliposis/

## ACKNOWLEDGEMENTS

Patients and their caregivers

## Footnotes

**Institutional review board statement**: The study received approval from the ethics committee of Puerta de Hierro University Hospital, internal code: PI_48/24.

**Informed consent statement**: written informed consent for data publication was obtained from patients.

