## Supplementary figures and images for "Optimizing Genetic Testing Strategy for Suspected Attenuated Adenomatous Polyposis: Effective Solutions in Public Health Systems"

### Supplementary Figure 1.

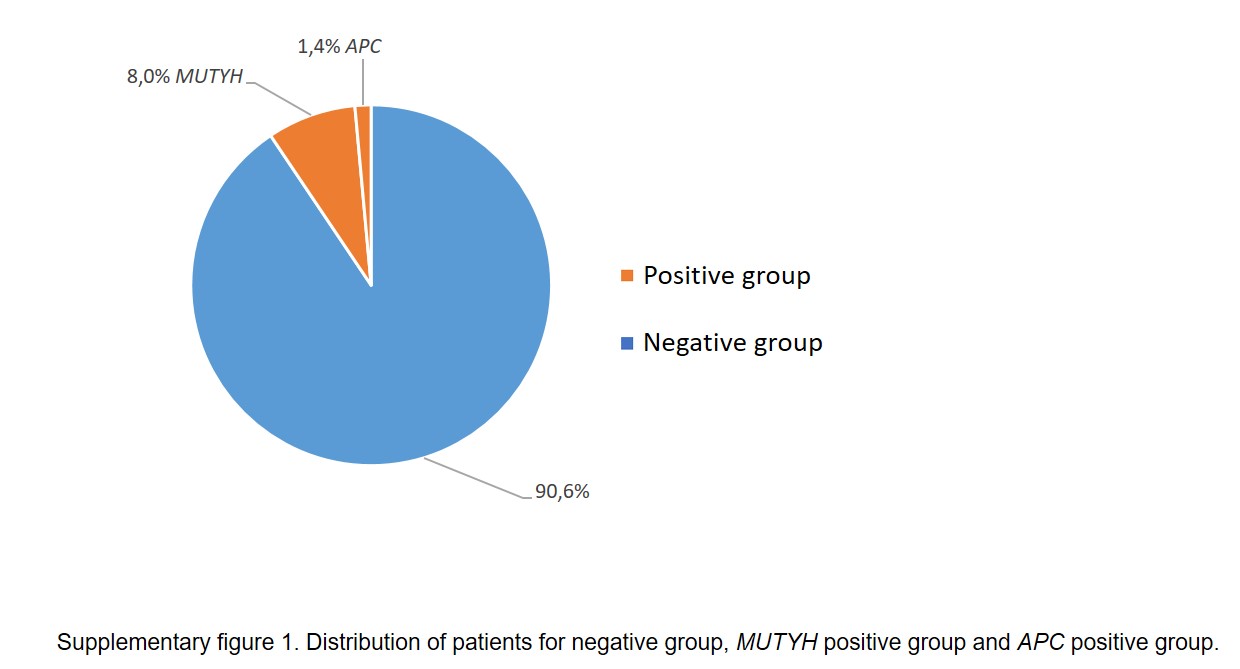

### Supplementary Figure 2.

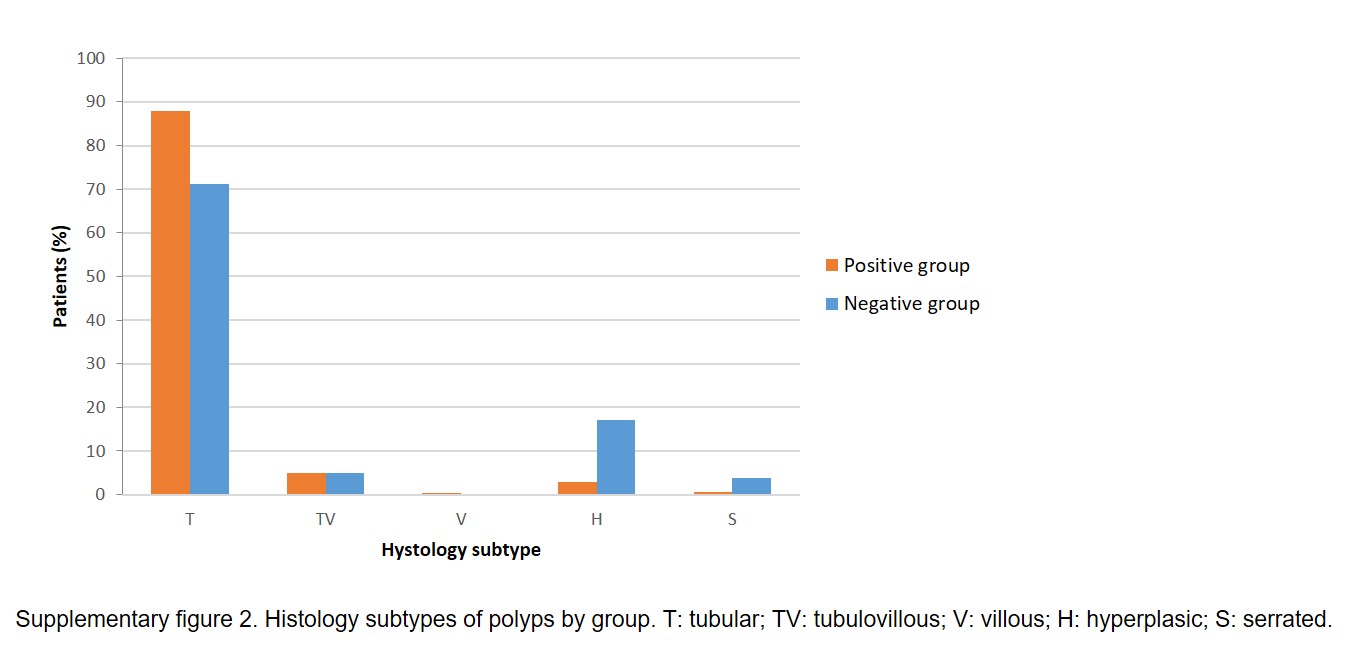
